# Dentate nucleus deep brain stimulation for spinocerebellar ataxia: results of a six-month follow-up

**DOI:** 10.1101/2025.02.10.25321730

**Authors:** Liang Zhao, Chang Qiu, Wenwen Dong, Bei Luo, Jian Sun, Jiuqi Yan, Xiang Wei, Guanghan Lu, Jingxuan Liu, Wenbin Zhang

## Abstract

Spinocerebellar ataxia (SCA) is a genetically heterogeneous neurodegenerative disorder lacking effective treatments currently. While noninvasive cerebellum neuromodulation showed positive results, invasive techniques like deep brain stimulation (DBS) have not been fully assessed for SCA patients. This study evaluated the treatment outcomes of DBS targeting the cerebellar dentate nucleus (DN) in six SCA patients (SCA1, SCA3, SCA12) over a six-month follow-up. Bilateral DN-DBS leads were precisely implanted using a neurosurgical robot. The stimulation parameters, including current, frequency, and pulse width, were programmed iteratively at predefined intervals, and the improvements in ataxia symptoms were evaluated based on the Scale for the Assessment and Rating of Ataxia (SARA) and the International Cooperative Ataxia Rating Scale (ICARS). Current results showed that current amplitude was fundamental to ensuring therapeutic efficacy, while frequency differentially alleviated tremor with high-frequency stimulation and gait disorder at low frequencies. At the six-month follow-up, the SARA and ICARS scores decreased by 43% (8.17 ± 2.58 vs. 14.33 ± 1.51, *p*=0.014) and 51% (18.67 ± 7.50 vs. 38.17 ± 8.13, *p*=0.013) respectively compared with baseline. Our study demonstrated the promising therapeutic benefits of DN-DBS for SCA patients and provided preliminary experience for individualized parameter programming.

## Introduction

Spinocerebellar ataxia (SCA) is a subtype of hereditary cerebellar ataxias and an autosomal dominant disorder characterized by progressive neurodegeneration ^1^. The main clinical signs are gradually worsening balance or coordination deficits and dysarthria. Epidemiological studies estimate a global SCA prevalence of 0 to 5.6 cases per 100,000 population, with significant geographical variations ^2^. SCAs present with significant genetic and phenotypic heterogeneity, with SCA3 as the most prevalent subtype, followed by SCA2 and SCA6 ^2^. Although not as intensively investigated to date, the mechanisms of SCA development may be related to polyglutamine toxicity, misfolded protein aggregation, ion channel dysfunction, and mitochondrial impairment ^3,4^.

Although no disease-modifying therapies have been approved so far, pioneering researches have revealed that neuromodulation therapies could be promising treatment approaches. Previous clinical trials have demonstrated the efficacy of repetitive transcranial magnetic stimulation (rTMS) and transcranial direct current stimulation (tDCS) targeting the cerebellum to reduce the ataxia severity in SCA patients ^5–7^. These noninvasive brain stimulation techniques have been proved to notably improve postural stability, ambulatory function, and limb coordination. These improvements can be quantified using the Scale for the Assessment and Rating of Ataxia (SARA) and the International Cooperative Ataxia Rating Scale (ICARS) ^8,9^. Besides TMS and tDCS, there is a lack of research on the treatment of SCA using invasive neuromodulation techniques in the SCA, such as deep brain stimulation (DBS). Cury et al. demonstrated that DBS of the cerebellar dentate nucleus (DN) improved cerebellar tremor in SCA patients ^10^. Since this study included only two SCA type 3 patients, further clinical research with more participants is required to validate the benefits of DN-DBS.

In this study, we included six SCA patients, specifically types SCA1, SCA3, and SCA12, who underwent DN-DBS surgery and were followed up for six months to observe improvements in ataxia. We also aimed to explore the potentially applicable range of stimulation parameters to assess the potential value of DN-DBS in treating SCA patients and to provide insights for stimulation programming.

## Methods

### Participants

Six patients with severe spinocerebellar ataxia were included. The inclusion criteria were as follows: 1. diagnosed with SCA confirmed by whole exome sequencing; 2. aged between 18 and 65 years; 3. chronic disease course over one year. The diagnoses of SCA types of the participants were evaluated by two experienced neurologists. Data were collected from December 2023 to January 2025. All patients provided informed consent. The study was approved by the ethics committee of the Affiliated Brain Hospital of Nanjing Medical University (ethics number: 2024-XJS006-02) and conducted according to the principles of the Declaration of Helsinki. Surgical procedure

On the day of surgery, computed tomography (CT) images were obtained after four or five bone-implanted markers were fixed. After that, CT images, pre-scanned three-dimensional T1-weighted, T2-weighted, and susceptibility-weighted magnetic resonance imaging (MRI) images were imported to the Reme-Studio (Remebot, Beijing, China) surgical plan system to accomplish image fusion, target planning, and implantation trajectory planning. Additionally, magnetic resonance venography images were utilized to visualize the occipital venous sinuses, facilitating the avoidance of hemorrhage during electrode implantation. The target for electrode implantation was set to ensure that as many contacts as possible were positioned within the DN. The registration error was confirmed to be below 0.3 mm for tracker-to-image and under 0.1 mm for tracker-to-robot alignment across all patients. Patients underwent implantation of DBS electrodes (PINS L301s, Beijing, China) bilaterally at the DN using the RM-100 neurosurgical robot (Remebot, Beijing, China) and an implantable pulse generator (IPG) under general anesthesia.

### Outcome Measures

The SARA and ICARS scales were used to assess the ataxia symptoms of the patients at baseline and follow-up. MRI (1.5T or 3.0T) scans, conducted one week post-surgery with the IPG remaining switched off, were utilized to determine the active electrode for initial programming. The initial stimulation parameters were set as 2.0 mA, 60 Hz, and 60 μs. During the first two months after the activation of the stimulator post-surgery, stimulation parameters, including current, pulse width, and frequency, were adjusted multiple times until improvements in SARA or ICARS scale scores were observed without significant adverse effects, thereby establishing a potential parameter range for each patient. Following this optimization phase, stimulation parameters were adjusted every two months. Assessment of the patients’ rating scales was performed by an independent physician who was not involved in adjusting the stimulation parameters. The Kruskal–Wallis test was applied for comparisons among more than two groups. Following the Kruskal–Wallis test, Dunn’s post hoc correction analyses were conducted to perform pairwise comparisons between groups. A two-tailed p-value <0.05 was considered statistically significant. All statistical analyses were performed using the R software (v4.3.3, www.r-project.org).

## Result

### Demographic and Clinical Characteristics

The baseline demographic and clinical features of the participants are summarized in Table 1. Four SCA1, one SCA3, and one SCA12 patient were included in the present study. Patient 1 is the uncle of patient 3 on the maternal side. Average baseline age was 51 ± 13.88 years, and average illness duration was 6 ± 3.43 years. Baseline mean scores were 14.33 ± 1.51 for SARA and 37 ± 8.69 for ICARS. All patients underwent cerebellar DBS surgery targeting the bilateral DN. Postoperative CT and MRI scans confirmed accurate electrode placement in all cases, with no instances of intracranial hemorrhage. One patient experienced moderate postoperative dizziness and vomiting due to a unilateral perielectrode edema, which resolved within several days as the edema subsided. All patients were followed up at the scheduled intervals, which included assessments using the SARA and ICARS scales, as well as optimization of stimulation parameters.

**Table 1.**
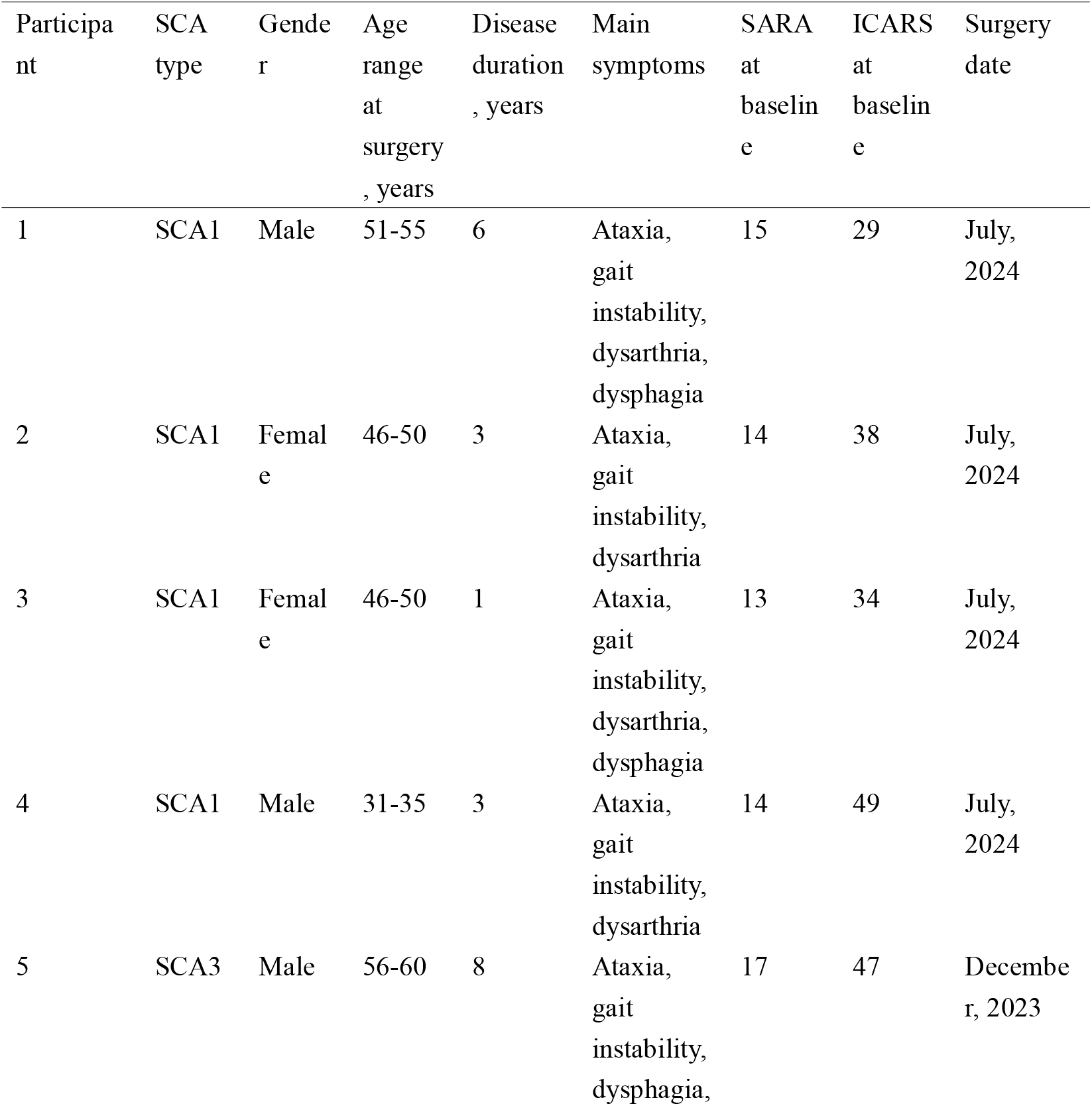
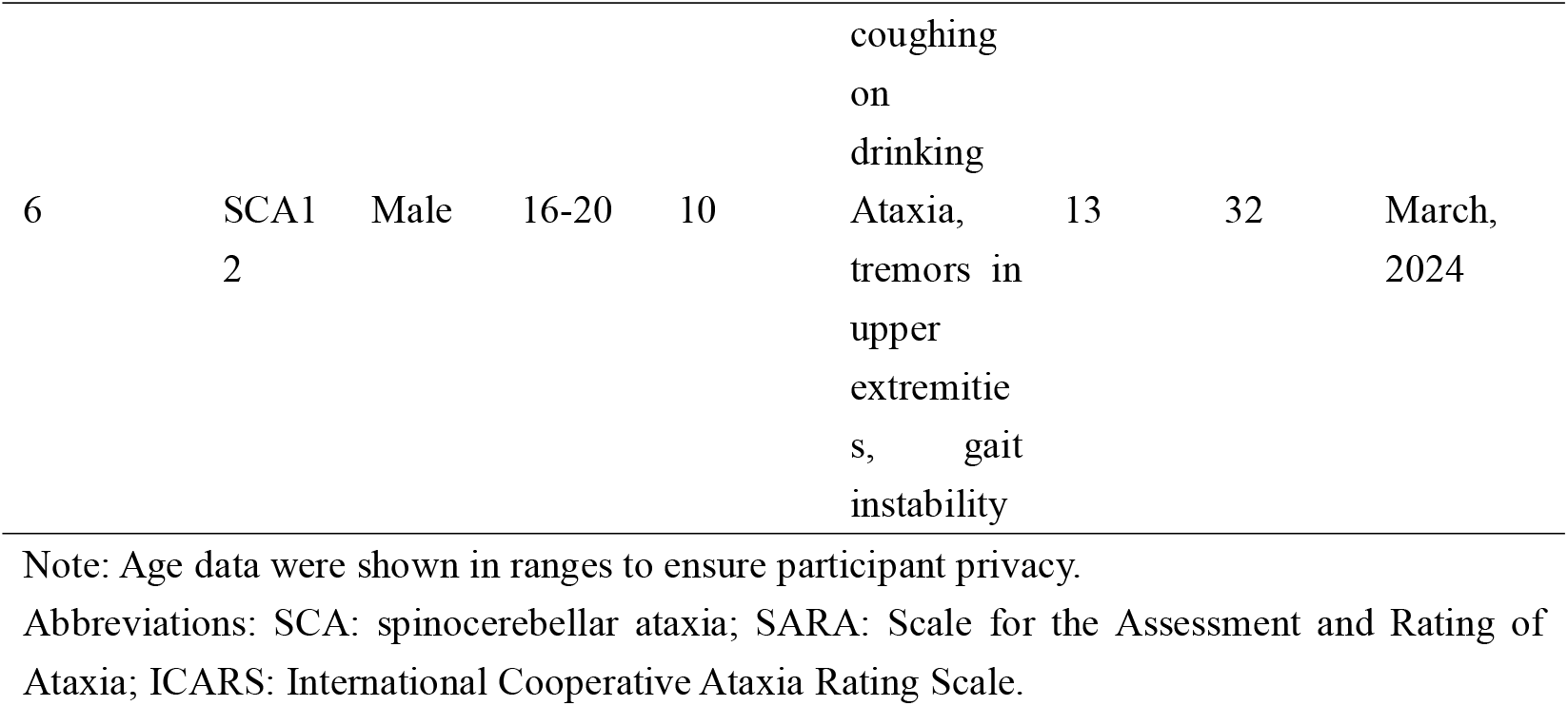
Clinical characteristics and baseline ataxia scaling scores of the six SCA patients.

### Patient outcomes

Based on postoperative cranial CT and MRI scans, a double monopolar stimulation mode was employed, utilizing the two contacts closest to the DN as the cathodes and the IPG case as the anode. The stimulation was initiated with conservative parameters (2.0 mA, 60 Hz, 60 μs), and no discomfort or adverse symptoms were reported by the patients during the initial programming phase. Following each adjustment of stimulation parameters, changes in SARA and ICARS scores typically emerged within days to weeks, accompanied by patient-reported subjective improvements, predominantly in gait stability and ataxia severity. Based on our experience with these six SCA patients, current amplitude exerted a predominant influence on symptom alleviation. However, excessive current amplitudes induced adverse effects such as dizziness and exacerbated gait instability. Pulse width appeared to modulate the therapeutic window: moderate increases improved motor symptoms, whereas excessive pulse widths triggered side effects analogous to those of high-current stimulation. Frequency adjustments differentially impacted tremor and gait patterns. Specifically, low-frequency stimulation enhanced gait performance, whereas high-frequency stimulation ameliorated tremor but was associated with worsened gait instability. The electrode configurations and stimulation parameters for the six SCA patients at six months post-DBS surgery are summarized in Table 2.

**Table 2.** Stimulation parameters and the improvements of ataxia scaling scores for SCA patients at the six-month follow-up.

| Participant | Active contacts (Left) | Active contacts (Right) | Frequency (Left/Right) | Pulse width (Left/Right) | Current amplitude (Left/Right) | % Changes in SARA to baseline | % Changes in ICARS to baseline |
| --- | --- | --- | --- | --- | --- | --- | --- |
| 1 | 1,2(-)/C(+) | 1,2(-)/C(+) | 40Hz | 100/100 μs | 4.6/4.6mA | 50 | 48.28 |
| 2 | 3,4(-)/C(+) | 3,4(-)/C(+) | 30Hz | 130/130 μs | 5.2/5.0mA | 42.86 | 60.53 |
| 3 | 1,2(-)/C(+) | 1,2(-)/C(+) | 40Hz | 130/130 μs | 5.2/5.5mA | 30.77 | 32.35 |
| 4 | 3,4(-)/C(+) | 1,2(-)/C(+) | 50Hz | 80/80 μs | 3.3/3.3mA | 39.29 | 69.39 |
| 5 | 2,3(-)/C(+) | 1,2(-)/C(+) | 40Hz | 100/100 μs | 4.2/4.2mA | 29.41 | 31.91 |
| 6 | 1,2(-)/C(+) | 2,3(-)/C(+) | 60Hz | 100/100 μs | 3.5/3.2mA | 69.23 | 62.5 |

Note: Electrode contacts are numbered sequentially from ventral to dorsal orientation, with contact 1 representing the most ventral position and contact 4 the most dorsal site.
Abbreviations: SARA: Scale for the Assessment and Rating of Ataxia; ICARS: International Cooperative Ataxia Rating Scale.

Stimulation parameter adjustments and bimonthly follow-up assessments revealed significant changes in both SARA (Kruskal-Wallis test, p=0.0038) and ICAR scores (p=0.0068). No significant reductions in SARA or ICARS scores were observed at the 2-month follow-up (all *p* > 0.05). Patients exhibited a significant reduction in SARA scores at 6 months post-stimulation initiation compared to baseline (8.17 ± 2.58 vs 14.33 ± 1.51, *p*=0.014), with a mean improvement of 43% from initial values (Fig. 1A). Concurrently, the ICARS scores (18.67 ± 7.50) exhibited a pronounced decrease from initial values (38.17 ± 8.13), achieving a 51% improvement with statistical significance (*p*=0.013) (Fig. 1B).

**Figure 1.**
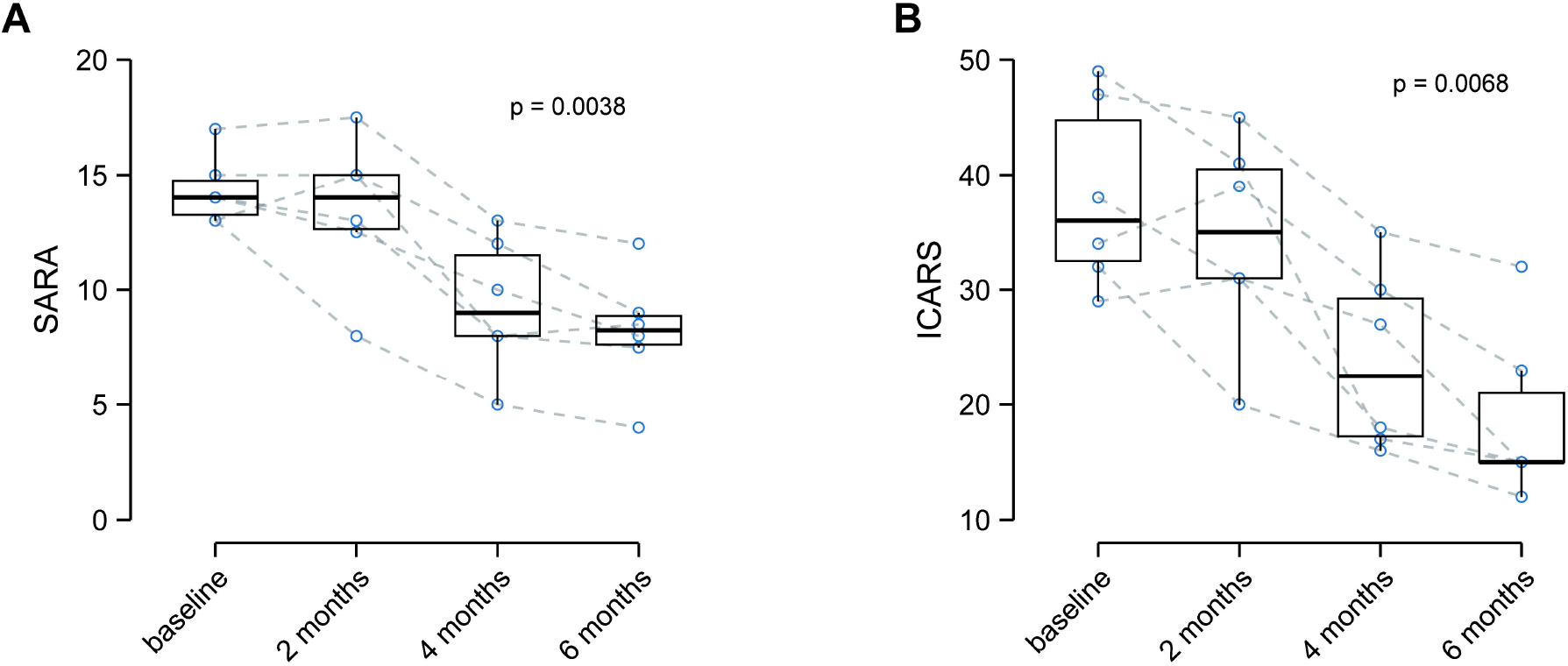
Changes in SARA and ICARS scores of six SCA patients during a six-month follow-up. Boxplots of SARA (**A**) and ICARS (**B**) scores for patients at each follow-up time point. The dashed lines represent the dynamic changes in scores for each patient. *P*-values were calculated using the Kruskal–Wallis test.

## Discussion

Previous studies have highlighted the important role of the DN in the interactions between the cerebellum and basal ganglia ^11,12^. . The DN connects with the cortical-cerebello-thalamocortical circuit to modulate motor and cognitive behaviors by receiving projections from the cerebellar cortex ^13^. Moreover, Hoover et al. discovered that DN functions as the origin of the dentate-thalamo-cortical (DTC) pathway to communicate the cerebellum with the primary motor cortex ^14^. Therefore, regulation of the output of DN and the afferents of the DTC pathway may change the excitability of M1 and further influence motor symtom. Previous clinical researches using noninvasive brain stimulation methods, including TMS and tDCS, have demonstrated that cerebellar stimulation can change the corticospinal or intracortical excitability and plasticity of the primary motor cortex ^15,16^. However, the stimulation of these studies broadly targeted the cerebellum without focusing on the DN, which limits elucidation of the DN’s functions in motor regulation. Cury et al. firstly reported that DN-DBS significantly reduced the tremor severity in five cerebellar ataxia patients, consisting of two SCA3, one cerebral palsy, and two cerebellar stroke patients ^10^. However, DN stimulation in the study failed to reduce the scaling score of ataxia, which is the main symptom of SCA patients. Moreover, a sample size of two patients included was insufficient to conduct a statistical analysis to evaluate the differences before and after DN-DBS for SCA patients.

Previously published works suggested that DN can be divided into two different regions based on neuroimaging techniques ^17,18^. The dorsorostral domain may target the lateral part of the ventral lateral (VL) nucleus and correlate with motor function, while the ventrocaudal portion may connect the medial part of VL and modulate the nonmotor symptoms. However, no study has elucidated the functions of the two subregions of DN in mediating movement disorders of SCA patients. Considering the target selection for electrode implantation is a key factor affecting the efficacy of DBS treatment, the sweet spots definition of DN in SCA patients should be further confirmed by large-scale clinical studies. Moreover, the question of whether the optimal stimulation targets for different SCA genetic types differ should be answered. The identification of the sweet spot may also facilitate the design of the electrode insertion trajectory and angle and enhance surgical safety by avoiding the sagittal sinus. For example, dorsolateral border stimulation of subthalamic nucleus (STN) was proved to be associated with reduced rigidity and akinesia and improvement of overall motor function in Parkinson’s disease ^19^. With respect to DN-DBS surgery, the cranial entry point of a steep medial-to-lateral trajectory may be too close to the sagittal sinus, while a lateral-to-medial approach may result in insufficient contacts placed within DN, which predicts poor efficacy of treatment. In our study, we implanted the electrodes to cover as many contacts as possible within DN and selected the active contacts which showed the most significant improvement in SARA or ICARS scores.

It is well established that stimulation parameters, including current/voltage, frequency, and pulse width, critically influence DBS efficacy across movement disorders. Our results showed that current/voltage amplitude still dominates therapeutic effects in DN-DBS analogous to stimulation programming in other movement disorders. In Parkinson’s disease, high-frequency stimulation typically improves most parkinsonian symptoms, while lower frequencies are implicated in axial signs control like gait impairment and speech ^20^. Similarly, our findings suggested that lower-frequency stimulation may stabilize gait in SCA patients, whereas higher frequencies could significantly alleviate tremor. These frequency-specific effects may reflect differential modulation of neural circuits within the DN-involved pathways. Pulse width adjustments likely modulate the spatial spread of stimulation in DN, partially explaining the side effects caused by excessive values like high-amplitude parameters presumably because. In this study, the non-significant SARA and ICARS improvements in the first two months of follow-up were presumably due to the initial conservative parameter settings to avoid unexpected side effects, which delayed therapeutic thresholds. Also, iterative parameter adjustments and neuroplastic adaptation may limit the immediate therapeutic effects, emphasizing the need for careful frequent assessment and individualized programming in the early phase ^21^. Variability in SCA genetic subtypes may require subtype-specific parameter optimization, which was not systematically explored in our present study.

Although our findings provided new evidence supporting the clinical value of DN-DBS for the treatment of SCA, we still have to acknowledge some limitations in the study. Firstly, the limited sample size and insufficient genetic variety make it difficult to draw solid conclusions on the generalizability of DN-DBS efficacy for SCA. Different types of SCAs harbor diverse repeat expansion and conventional mutations, as well as heterogeneous clinical phenotypes, which may differentially respond to DN-DBS ^22^. Secondly, the underlying mechanisms of DN stimulation in SCA have not been fully investigated. Numerous studies have used radiomics, connectomics, and electrophysiological techniques to examine the neuromodulation processes of DBS in movement disorders such as Parkinson’s disease and dystonia ^23,24^. More extensive works are now required to clarify the mechanisms and identify potential biomarkers using multimodal approaches. Directional lead composed of segmented electrodes can generate a stimulation field in a particular direction, which greatly enhances the personalization and flexibility of modulation ^25^. It is promising that directional lead may play an important role in exploring the functional recognition of subregions within the DN and stimulation patterns for SCA. However, considering the high costs of directional leads, all leads used in the present study were conventional electrodes.

In summary, our study showed the encouraging therapeutic benefits of DN-DBS for six patients with SCA, showing significant reductions in SARA and ICARS at long term follow-up. Moreover, our results provided preliminary experience for DN-DBS parameter programming for SCA patients, which may have important implications for strategies to individualize treatment.

## Acknowledgments

We thank all patients for their study participation. This work was supported by the Special Funds of the Jiangsu Provincial Key Research and Development Projects (Grant No.: BE2022049, BE2022049-1) and High-end Medical Equipment Promotion and Application Program (Grant No.: 2024TGYY46).

## Data Availability

Data and further information used in the current study are available from the corresponding author on reasonable request.

